# Evaluation Domains and Measurement Approaches for Assessing the Performance of China’s Family Doctor Contract Services: A Scoping Review Protocol

**DOI:** 10.1101/2024.09.20.24314035

**Authors:** Yang Wang, Hua Jin, Dehua Yu

## Abstract

**Background:** Family doctor contract services (FDCS) in mainland China are designed to foster long-term relationships between family doctors and residents, promoting primary care delivery through teamwork. By 2035, coverage is expected to exceed 75%. Following the release of national standardized guidelines in 2018, the policy focus has shifted from expanding coverage to improving service quality. However, the evaluation domains and measurement approaches currently used to assess FDCS remain fragmented and lack a consensus-based standard. This scoping review aims to systematically map the full spectrum of evaluation domains and analyze the measurement approaches used to assess the performance of FDCS in mainland China.

**Methods:** Conducted in accordance with JBI methodology, we will search CNKI, Wanfang, PubMed, Web of Science, Embase, and Google Scholar for literature published from January 2019 onwards. This timeframe specifically accounts for the publication lag following the 2018 milestone national policy. Sources will include policy documents, quantitative studies, and qualitative studies. A comprehensive full-text data extraction strategy will be employed for all included studies. We will categorize all identified evaluation domains using the Donabedian (Structure, Process, Outcome) framework. Results will be synthesized through a two-stage strategy: (1) mapping the macro-alignment of evaluation domains across the Structure-Process-Outcome continuum by evidence source (Sankey Diagram), and (2) cataloging the specific characteristics of Outcome-related measurement tools, including target constructs, number of items, and reported psychometric properties (Instrument Inventory Table).

**Discussion:** This review will provide the first comprehensive map of the FDCS evaluation ecosystem during its standardized development phase. It will objectively identify alignments and gaps between policy mandates (intended evaluation domains), research practices (actual measurement approaches), and stakeholder perspectives. By mapping this landscape, the review aims to provide a clear evidence base for future evaluations and the development of a consensus-based core outcome set.

**Systematic review registration:** Registered in OSF on September 20, 2024 (osf.io/z3eju)

## Introduction

The “family doctor contract service (FDCS)” implemented in mainland China is built upon long-term doctor-patient and social relationships established between family doctors and contracted residents. It supports family doctors in providing primary care services to community and rural residents through teamwork and patient-doctor collaboration^1,2^. According to the goals set by the National Health Commission’s Primary Healthcare Department, by 2035, the coverage rate of family doctor contract services in China is expected to exceed 75%^3^. This suggests that family doctor contract services are gradually becoming the main approach for primary care facilities to deliver primary care and essential public health services to community residents.

As summarized by Stange et al., the purpose of using metrics to assess health service quality is to guide actions and highlight important issues that may be overlooked in routine work^4^. In Donabedian’s classic theoretical model, three key elements—”structure,” “process,” and “outcome”—are prioritized when assessing healthcare service quality. Among these, “outcome” is not only the ultimate standard for verifying the effectiveness and quality of health services but is also prone to measurement bias due to variations in “time,” “environment,” “structure,” and “process”^5^. Therefore, measuring “outcome,” as the most fundamental and critical aspect of healthcare quality evaluation, must involve the careful selection of dimensions and tools to ensure the results are accurate, relevant, and easy to use, thereby providing meaningful support for future quality improvements.

However, despite the policy transition towards high-quality development since 2018, there remains a lack of comprehensive understanding regarding the specific domains and measurement approaches used to evaluate FDCS. Recent key policy documents and government surveys ^6-9^ highlight a mixture of performance and outcome indicators, including “health improvement,” “control of health risk factors,” “number of visits,” “patients’ choice of primary care institution,” “patient satisfaction,” “service quality,” and “coverage rate.” Overall, the indicators presented across these documents span different methodological levels (structure, process, and outcome) without a unifying framework, resulting in an evaluation landscape that is diverse, fragmented, and somewhat disorganized.

Research in this field also shows similar characteristics. According to the literature, the dimensions and indicators used to measure the performance of family doctor contract services have included at least ten different outcome domains, such as “health”^10-12^, “functional features”^13^, “chronic disease control”^14^, “patient loyalty”^15^, “doctor-patient trust”^16-18^, “resident contract rate”^19^, “service utilization” ^20,21^, “patient satisfaction”^22^, “residents’ medical costs”^23^, and “receipt of chronic disease and preventive medicine screenings”^24,25^. Moreover, different studies employ various tools to assess the same outcome dimension, which complicates the comparison and aggregation of findings, further contributing to the fragmented nature of research in this area. For example, tools for measuring “health” include a 5-point Likert scale^10^, the EQ-5D-5L scale^11^, and the SF-36 scale^12^, each reflecting different core constructs.

This documented fragmentation in both policy and research highlights an urgent need for a comprehensive map of the existing evaluation landscape. Without a clear synthesis of what is being measured and how, it is impossible to identify gaps, standardize approaches, or develop a consensus-based core outcome set.

The aim of this scoping review is to comprehensively summarize the evaluation dimensions and measurement approaches previously used to assess the performance of family doctor contract services in mainland China, focusing on the standardized development phase (literature published from 2019 onwards, accounting for the publication lag of the 2018 policy).

Following an initial search of PROSPERO, the Cochrane Database of Systematic Reviews, JBI Evidence Synthesis, and other key databases, we identified a recently registered mixed-methods systematic review regarding FDCS (PROSPERO CRD42024587272) ^26^. However, that review aims to synthesize the clinical and implementation effectiveness (specifically focusing on chronic disease management outcomes, facilitators, and barriers). In contrast, our scoping review focuses strictly on the evaluation methodology itself—mapping the entire spectrum of domains (spanning Structure, Process, and Outcome) and triangulating policy mandates with actual measurement practices. Therefore, the objective of our review is entirely distinct, uniquely serving as a methodological foundation for the future development of a consensus-based core outcome set (COS).

### Review question

What evaluation domains and measurement approaches have been used to assess the performance of family doctor contract services in mainland China since its standardized development phase (2019 onwards)?

In this review, “Evaluation Domains” refer to the specific content areas being assessed (e.g., blood pressure control, patient satisfaction, service accessibility, provider workload), which will be analyzed through the theoretical lenses of the Donabedian model (Structure-Process-Outcome).

“Measurement Approaches” are defined as the specific methodologies used to capture data, encompassing specific indicators, measurement tools (e.g., scales, questionnaires), target constructs, number of items, and data sources/respondents (e.g., patient-reported, provider-reported, administrative records).

“Performance” is operationalized as encompassing the Structure, Process, and Outcome dimensions of the Donabedian model.

### Inclusion criteria

#### Participants

Stakeholders involved in China’s family doctor contract services (including contracted residents, family doctors, and primary care managers/teams).

#### Concept

Evaluation dimensions (structure, process, and outcome) and measurement tools used to assess FDCS performance.

#### Context

China’s primary care context.

#### Types of sources

The included literature must fall into one of the following three categories: government-issued policy documents, quantitative studies, and qualitative studies.

## Methods

The proposed scoping review will be conducted in accordance with the JBI methodology for scoping reviews^27^. This review is based on the present protocol. Any deviations from the protocol will be reported in the relevant section of the methods, along with a justification.

### Inclusion Criteria

1. The included literature must fall into one of the following three categories: government-issued policy documents, quantitative studies, and qualitative studies.
2. Each type of literature must meet the following criteria:
  (1) Policy documents: The document must focus on family doctor contract services and explicitly mention the assessment and evaluation of the quality or outcomes of these services.
  (2) Quantitative studies: The study population must consist of residents contracted with family doctors, meaning individuals who have signed a service agreement with family doctors in China. The exposure or intervention must involve family doctor contract services or specific components of these services, and the study must report measurable indicators related to the performance, quality, structure, process, or outcomes resulting from the exposure/intervention.
  (3) Qualitative studies: The phenomenon of interest must be the stakeholder perceptions, experiences, or valued aspects related to the structure, process (e.g., quality), or outcomes of family doctor contract services.
3. Only literature published from January 1, 2019, onwards will be included. This timeframe captures the standardized and high-quality development phase following the milestone 2018 National Guidelines^8^.
4. Both quantitative and qualitative studies can include published literature or gray literature, such as graduate theses.
5. Literature published in English and Chinese languages will be included.

### Exclusion Criteria

1. We will strictly exclude opinion pieces, editorials, daily news bulletins, and narrative reviews that lack a formalized methodology or structured empirical data.
2. Studies exploring factors influencing residents’ intentions to contract, rather than evaluating the actual performance or outcomes of the services, will be excluded. Similarly, documents focusing broadly on “primary health care” without specifically examining FDCS will be excluded.
3. Studies evaluating specific clinical technologies or pharmaceutical interventions (e.g., drug efficacy trials) that operate independently of the routine FDCS management model (as defined by the 2018 Guiding Opinions^8^) will be excluded.
4. Studies focused on contexts outside of mainland China (e.g., Hong Kong, Macau, or Taiwan) will be excluded.
5. We will exclude systematic reviews, other scoping reviews, and incomplete sources published as ‘abstracts only’ (e.g., conference abstracts) that do not provide sufficient methodological data for full-text extraction.

### Search strategy

The search strategy will be divided into two tracks based on the type of evidence:

### Track 1: Government-issued policy documents

We will systematically search the official websites of the National Health Commission of the People’s Republic of China and the 31 provincial-level administrative health commissions. The search will use key terms and their policy-related synonyms (e.g., “family doctor contract service”, “signing service package”). To ensure transparency, a dedicated Policy Document Selection Flowchart will be constructed to record the screening and inclusion process, independent of the academic literature PRISMA-ScR flow.

### Track 2: Quantitative and qualitative studies

We will develop two search strategies (English and Chinese) based on the JBI scoping review methodology^27^, focusing on literature published from 2019 onwards.

For the English-language literature, an initial search was conducted in PubMed and Embase. Following frequency analysis using VOSviewer (developed by the Centre for Science and Technology Studies, CWTS, at Leiden University, Netherlands), a comprehensive search strategy was developed (see Appendix 1). This strategy is designed to be highly sensitive, combining key concepts related to both “family doctor” and “general practitioner” with terms related to “contract” or “signing” to identify all relevant studies. This strategy will be applied in a second round of searches in PubMed, Embase, Web of Science, and Google Scholar (screening the first 200 relevant results to ensure reproducibility). Finally, the reference lists of all relevant articles will be reviewed to identify additional sources.

For the Chinese-language literature, an initial search was conducted in the CNKI and Wanfang databases. Based on the JBI methodology for triangulating diverse sources, the final search strategy (see Appendix 1) combines the core concept “family doctor contract service” with its key policy-related synonyms. Given the unique indexing characteristics of Chinese databases—which aggregate a massive volume of non-academic government news, daily bulletins, and unformatted commentaries alongside peer-reviewed literature—a targeted syntax condition (AND “research” OR “methods” in Abstract/Keywords) was applied. This serves as a necessary and methodologically sound pre-screening filter to isolate formal empirical studies from administrative noise. A second round of searches will be conducted in CNKI and Wanfang, followed by a rigorous review of reference lists (snowballing) to mitigate any risk of inadvertently excluding non-standard qualitative studies.

Given that the primary sources of academic gray literature in mainland China (such as master’s and doctoral dissertations) are comprehensively indexed within the CNKI and Wanfang databases, these platforms will serve as our designated gray literature repositories alongside official government websites.

### Study selection

After the search, all information regarding the identified articles will be imported into EndNote X9.2 software (Clarivate Analytics, Philadelphia, USA, 2019), where duplicates will be removed. The study selection will then be performed by importing the literature information into Rayyan, a web and mobile app designed for conducting systematic reviews.

Two reviewers familiar with the topic (YW, HJ) will independently screen the titles, keywords, and abstracts of the first 50 references as a pilot test to refine the eligibility criteria and ensure a consistent understanding. Weekly comparisons and group discussions will be conducted. Full screening will begin once inter-rater reliability reaches an acceptable threshold (e.g., >90% agreement). If the two reviewers fail to reach a consensus during the screening process, a third reviewer (DY) will moderate discussions to resolve discrepancies.

The study selection process will be illustrated using a PRISMA-Scr flow diagram, detailing the stages from the initial search, duplicates removed, title and abstract screening, full-text retrieval, and any additional sources identified through reference lists. Details of excluded studies and the reasons for their exclusion at the full-text stage will be recorded and securely stored. Government policy documents were selected separately based on relevance to FDCS evaluation standards. These documents are not subject to the PRISMA flow but are listed as a supplementary data source for the multi-source comparative mapping.

### Data extraction

The data extraction for this review will be conducted using three distinct extraction forms, tailored to the specific nature of each evidence type (policy, qualitative, and quantitative) to facilitate our multi-source comparative mapping.

1. For government-issued policy documents, the data extraction will include: issuing agency, year of publication, region/jurisdiction (e.g., national level, or specific province/city), and the specific evaluation dimensions or indicators mandated (e.g., ‘contract rate’, ‘team configuration’). Each extracted indicator will be classified by the review team according to the Donabedian (Structure, Process, Outcome) framework.
2. For qualitative studies: the data extraction will focus on: author, year, participant type, participant location/setting (e.g., Urban Shanghai, Rural Guizhou), data collection method, themes and sub-themes related to the performance or outcomes of FDCS, conceptual interpretation (narrative definition or illustrative quotations from the original text), and classification according to the Donabedian framework.
3. For quantitative studies, a comprehensive full-text extraction will be conducted for all included articles. The data extraction will capture: (a) Study characteristics: author, year, province, population, study design; (b) Evaluation domains: mapped to the Donabedian framework (Structure, Process, Outcome). Where applicable, Outcome domains will be tagged with value-orientation labels (e.g., population health, patient experience, cost, equity) to facilitate the subsequent narrative discussion; and (c) Detailed measurement approaches: specific metric definitions, exact names of the measurement tools used (e.g., standardized scales, custom questionnaires, or administrative indicators), target constructs, number of items, reported psychometric properties (e.g., validity, reliability), and data sources/respondents (e.g., patient-reported, provider-reported, administrative records).

To ensure inter-rater reliability, the three extraction forms will be piloted by two reviewers (YW and HJ) on a purposive sample of 15–20 sources, distributed evenly across the three evidence types. This pilot phase will specifically focus on calibrating the complex quantitative classification criteria (S-P-O framework) and the ‘Data Source’ definitions through consensus meetings involving the third reviewer (DY). To minimize subjective classification bias, a pre-defined coding manual with formal decision rules for boundary cases (e.g., distinguishing a process indicator from an outcome indicator) will be iteratively refined during this phase. Any discrepancies will be resolved through discussion, and the extraction forms will be refined iteratively to maximize clarity. All extracted data will be securely stored with clear audit trails for tracking and reference.

### Data analysis and presentation

In this scoping review, data will undergo a multi-stage comparative mapping analysis. The analysis is designed to systematically map the findings and identify gaps within the evaluation ecosystem.

### Step 1: Evaluation Domain Mapping (Sankey Diagram)

We will synthesize the evaluation dimensions identified from all three evidence tracks (policy documents, quantitative studies, and qualitative studies). This will be visually presented using a Sankey Diagram with three pillars: ‘Evidence Source Type’ (Pillar 1), ‘S-P-O Category’ (Pillar 2), and ‘Specific Evaluation Sub-domains’ (Pillar 3). To prevent the mixing of conceptual granularities in Pillar3, extracted dimensions will be standardized using a hierarchical taxonomy through qualitative clustering, categorizing them into broad domains (e.g., ‘Clinical Quality’, ‘Experiential Metrics’) and specific sub-domains (e.g., ‘Blood Pressure Control’, ‘Satisfaction’). This single visualization will reveal which evaluation domains are prioritized, neglected, or misaligned across different evidence sources.

### Step 2: Measurement Instrument Inventory (Instrument Table)

Focusing on the subset of dimensions classified as “Outcome,” we will construct a comprehensive Measurement Instrument Inventory based on the full-text extraction of all included quantitative studies. This table will catalogue: the specific tools used (e.g., SF-36, EQ-5D-5L, custom questionnaires, or administrative indicators), their target constructs, the number of items, reported psychometric properties (e.g., internal consistency, structural validity), and corresponding data sources/respondents.

### Step 3: Narrative Synthesis

A descriptive narrative will synthesize the findings from Step 1 and Step 2, organized around two analytical foci:

1. Evaluation domain landscape: Drawing on the Sankey Diagram (Step 1), we will analyze the frequency and distribution of evaluation sub-domains across the three evidence sources (policy, quantitative, qualitative) and SPO categories. The narrative will identify the dominant measurement trajectories (i.e., which stakeholders measure what, and through which methodological pathways), as well as systematically under-represented domains.
2. Measurement instrument patterns: Drawing on the Instrument Inventory (Step 2), we will identify the convergence or divergence in tool selection for Outcome measurement, examining which instruments are most frequently adopted, whether standardized or custom-developed tools predominate, and what data sources (e.g., patient-reported, administrative) are most commonly used.

## Data Availability

All data produced in the present work are contained in the manuscript.

https://osf.io/z3eju

## Acknowledgements

Nil

## Funding

This research was funded by and the 2024 Shanghai City Yangpu District Science and Technology and Economic Committee Deepening Medical Reform Innovation Research Project (Grant No: YPYG202403); Shanghai Municipal Health Commission Health Policy Research Project (Grant No. 2023HP28&2023HP71), Shanghai Leading Talents Program (Grant No. YDH-20170627), and Discipline Leader Advancement Program of Yangpu District Central Hospital (Ye2202103).

## Ethics approval and consent to participate

Not applicable. This scoping review relies exclusively on the analysis of publicly available literature and government policy documents; thus, ethical approval was not required.

## Declarations

The authors declare no conflict of interest.

## Author contributions

Conceptualization, Y.W.; Methodology, Y.W and H.J; Data curation, Y.W and H.J; Formal analysis, Y.W.; Funding acquisition, H.J and D.Y; Project administration, D.Y; Resources, Y.W and H.J; Supervision, D.Y; Validation, Y.W; Writing—original draft, Y.W.; Writing—review and editing, Y.W., H.J., D.Y.. All authors have read and agreed to the published version of the manuscript.

## Appendices

### Appendix I: Search strategy

The search strategy is designed to be highly sensitive to capture all three evidence types (policy, quantitative, and qualitative) in accordance with JBI methodology.

#### Policy Documents

We will search the official websites of the National Health Commission of China and the 31 provincial-level administrative units in mainland China. The search will use a combination of key terms and their policy-related synonyms, including:

“家庭医生签约” (family doctor contract)

“签约服务” (signing service)

“全科医生签约” (general practitioner contract)

“家庭医生服务团队” (family doctor service team)

“签约服务包” (contract service package)

#### Quantitative/Qualitative Studies (English)

The search strategy will be adapted for each database. The following strategy for PubMed will serve as the foundation:

##### PubMed

((((“family doctor*”[Title/Abstract] OR “general practitioner*”[Title/Abstract]) AND (“contract*”[Title/Abstract] OR “sign*”[Title/Abstract] OR “signing”[Title/Abstract] OR “agreement*”[Title/Abstract])) OR “family doctor contract service*”[Title/Abstract])) AND (“China”[MeSH Terms] OR “China”[Title/Abstract] OR “Chinese”[Title/Abstract])

(345)

##### Embase

((((‘family doctor*’ OR ‘general practitioner*’):ti,ab,kw) AND ((‘contract*’ OR ‘sign*’ OR ‘signing’ OR ‘agreement*’):ti,ab,kw)) OR (‘family doctor contract service*’:ti,ab,kw)) AND ((‘China’/exp OR ‘China’:ti,ab,kw))

(643)

##### Web of Science (WOS)

(TI=(((“family doctor*” OR “general practitioner*”) AND (contract* OR sign* OR signing OR agreement*)) OR “family doctor contract service*”) OR AB=(((“family doctor*” OR “general practitioner*”) AND (contract* OR sign* OR signing OR agreement*)) OR “family doctor contract service*”)) AND (TI=(China OR Chinese) OR AB=(China OR Chinese))

(343)

##### Google Scholar

“Family doctor contract*” OR “GP contract*” OR “general practitioner signing” AND “China”

(500)

#### Quantitative/Qualitative Studies (Chinese)

##### China National Knowledge Infrastructure (CNKI)

(TKA=(“家庭医生签约”) OR TKA=(“全科医生签约”) OR TKA=(“家庭医生责任制”) OR TKA=(“签约服务包”) OR TKA=(“签约居民”) OR TKA=(“家庭医生服务团队”)) AND (TKA=(“研究”) OR AB=(“方法”) OR AB=(“调查”) OR AB=(“访谈”))

限定类别为”研究论文”

(1857)

##### Wanfang Data

题名或关键词:(“家庭医生签约” OR “全科医生签约” OR “家庭医生责任制” OR “签约服务包” OR “签约居民” OR “家庭医生服务团队”) AND (题名或关键词:(“研究”) OR 摘要:(“方法”) OR 摘要=(“调查”) OR 摘要=(“访谈”))

(2494)

Search conducted on: 18, November, 2025

## Notes

### Competing Interest Statement

The authors have declared no competing interest.

### Summary of Updates

We have submitted a revised version because a subsequent literature search revealed several technical oversights in the original plan that could have compromised our objectives. Accordingly, we have modified and further refined the approach.

